# The Prognostic Value of Normal Troponin on Admission in STEMI Patients: An 8-Year Multicenter Cohort

**DOI:** 10.64898/2026.07.21.26358627

**Authors:** Ohad Stoler, Raz Croitoru, Gassan Moady, Ofer Kobo, Zafrir Or, Mahmod Hammoud, Supriya Shore, Ariel Roguin, Idit Dobrecky Mery, Edo Y. Birati

## Abstract

**Background:** ST-elevation myocardial infarction (STEMI) remains a major cause of global mortality. While troponin is the gold-standard biomarker for myocardial injury, a subset of patients presents with troponin levels below the clinical “rule-in” threshold upon hospital admission. The long-term prognostic significance of these initial “low-troponin” presentations in a large-scale population remains insufficiently characterized.

**Methods:** We conducted a retrospective multicenter cohort study using the “KINERET” database, analyzing 8,394 patients diagnosed with STEMI who underwent percutaneous coronary intervention (PCI) at four academic medical centers in Israel between 2016 and 2023. Patients were stratified into two groups based on ESC rule-in criteria for high-sensitivity cardiac troponin (hs-cTn) at admission: a High trop group (above rule-in cutoff) and a Low trop group (below rule-in cutoff). The primary outcome was all-cause mortality at 5 years.

**Results:** Of the 8,394 patients (mean age 68.3±13.3 years; 76% male), 36.5% (n=3,064) presented with troponin levels below the rule-in cutoff. Patients in the High trop group were older and had a higher prevalence of comorbidities, including heart failure (46.7% vs. 28.7%) and chronic kidney disease (13.9% vs. 9%). The Low trop group demonstrated significantly higher survival rates at both 1 year (92.9% vs. 84.0%, p<0.001) and 5 years (85.7% vs. 75.0%, p<0.001). After adjusting for age, sex, and comorbidities in a multivariate Cox regression model, initially elevated troponin remained a robust independent predictor of 5-year mortality (HR 1.15, 95% CI 1.14-1.16, p<0.001), alongside age >75, female sex, and chronic kidney disease.

**Conclusions:** STEMI patients presenting with initial troponin levels below the diagnostic rule-in threshold have a significantly better short- and long-term prognosis compared to those with early troponin elevation. Moreover, admission troponin levels serve as a powerful predictor of 5-year mortality and may be used as an independent prognostic factor following STEMI.

## Introduction

Ischemic heart disease (IHD) remains a leading cause of mortality in the Western world, accounting for approximately 9 million deaths annually^1^. Myocardial infarction is currently classified based on the electrocardiographic (ECG) changes in the ST segment, specifically as ST-elevation myocardial infarction (STEMI) and non-ST-elevation myocardial infarction (NSTEMI)^2^.

Troponin is a protein found primarily in muscle cells and exists in several isoforms; troponin I and T are specific to cardiac muscle. An elevation of this protein indicates myocardial injury and begins within minutes of impaired blood supply. However, protein levels generally become measurable in the blood approximately 60 minutes after the onset of the event^3^. Several studies have demonstrated the prognostic importance of the troponin amplitude in STEMI patients^4^.

Many of the patients presenting with STEMI have normal troponin levels on admission. In this study, we aimed to evaluate the unique clinical characteristics of this patient population and its short- and long-term survival. We hypothesized that these patients may have more favorable outcomes than those with elevated troponin levels on admission.

## Methods

In the current study, we compared the outcomes of STEMI patients with normal high-sensitive cardiac troponin (hs-cTn) levels (below the rule-in for MI cut-off) on admission with those with elevated hs-cTn values (above the rule-in for MI cut-off), in four academic cardiac centers in Israel between 2016 and 2024, providing insights into their survival and clinical trajectory.

### Study Design and Data Source

This retrospective cohort study was conducted using data from the Israeli Ministry of Health database (“KINERET”). The “KINERET” database comprises clinical and demographic information on patients admitted to federal academic medical centers across Israel and provides a secure, cloud-based environment for data extraction and analysis. This infrastructure enables access to comprehensive electronic medical records while adhering to strict data security and privacy standards, using the Observational Medical Outcomes Partnership (OMOP) Common Data Model. Mortality data were obtained from the hospital’s records and cross-referenced with the Israeli National Population Authority to ensure completeness of follow-up.

### Study Population

The current study utilizes the medical records of patients hospitalized at the cardiovascular divisions of four academic medical centers (Tzafon Medical Center, Bnai Zion Medical Center, Galilee Medical Center, Hillel Yaffe Medical Center) in Israel between 2016 and 2023. Information such as medical history and laboratory results was retrieved from electronic medical records. The patient population included individuals diagnosed with myocardial infarction (MI), identified using ICD-9 codes 410.0-410.9 (without 410.7X; which is used mostly to define NSTEMI event), who underwent percutaneous coronary intervention (PCI) with ICD-9 codes 36.01, 36.06, 36.07, 36.09, 00.66, and 00.63.

### Troponin Cut-off criteria

Lithium-heparin anticoagulant-containing tubes were used to collect blood samples. Prior to the PCI, blood samples were taken for hs-cTn measurements. The cut-off troponin levels were determined by the European Society of Cardiology (ESC) rule-in criteria for MI^5^, in accordance with the specific hs-cTn in each medical center [52 ng/ml for hs-cTnt (Roche) and 64 ng/ml for hs-cTni (Abbott)].

### Outcome Assessment

The primary outcome was all-cause mortality at 5 years following the index MI. Secondary outcomes included all-cause mortality at 1 year and length of stay.

### Statistical Analysis

Statistical and data analysis were accomplished using Python version 3.9.7

Continuous variables are presented as mean ± standard deviation or median with interquartile range, as appropriate, and were compared using Student’s t-test or Mann–Whitney U test. Categorical variables are presented as counts and percentages and were compared using the χ^2^ test. Long-term survival was assessed using Kaplan–Meier analysis, with differences between groups evaluated using the log-rank test. Kaplan–Meier curves were generated to compare survival according to initial troponin levels.

To identify independent predictors of 5-year mortality, a multivariable Cox regression model was constructed, including clinically relevant covariates and variables associated with mortality in univariable analyses. Results are reported as Hazard ratios (HRs) with 95% confidence intervals (CIs).

### Ethical Approval

This study was conducted in accordance with the ethical principles of the Declaration of Helsinki and was approved by all four medical centers’ Helsinki committees.

## Results

A total of 8394 patients were included in the study. Among the study population, 6389 were male (76%). Patient ages ranged from 20 to 100 years, with a mean age of 68.3±13.3. Baseline characteristics are presented in Table 1. During hospitalization, the initial hs-cTn was elevated in 63.5% of patients (High trop group; n=5330) above the rule-in cutoff. 36.5% of patients had hs-cTn below the rule-in cutoff (Low trop group; n=3064). Patients in the High trop group were older, with an average age of 69.3 ± 13.7 years, compared with 66.6 ± 12.5 years in the Low trop group (p<0.001). A significant variation in gender distribution was also observed (p<0.001), with a greater proportion of males in the Low trop group (79.6%) compared to the High trop group (73.9%). Most comorbidities were significantly more prominent in the High trop group. Specifically, the High trop group had higher rates of prior diagnosis of heart failure (46.7% vs. 28.7%), chronic kidney disease (CKD; 13.9% vs. 9%), and diabetes (42.6% vs. 38.9%) compared with the Low trop group (p<0.001 for all; Table 1).

**Table 1.** Baseline Characteristics.

| Table 1 - Baseline Characteristics |  |  |  |  |
| --- | --- | --- | --- | --- |
| Characteristic<br>(count, %) | Trop > Rule In, No.<br>(%) n=5330 | Trop < Rule In, No.<br>(%) n=3064 | Total, No.<br>(%) n=8394 | p-value |
| Average Age (±SD) | 69.3 ± 13.7 | 66.6 ± 12.5 | 68.3 ± 13.3 | <0.001 |
| Male | 3949 (73.9) | 2440 (79.6) | 6389 (76.0) | <0.001 |
| Atrial fibrillation | 865 (16.2) | 454 (14.8) | 1319 (15.7) | 0.100 |
| Diabetes Mellitus | 2277 (42.6) | 1192 (38.9) | 3469 (41.2) | <0.001 |
| Hypertension | 3566 (66.7) | 1961 (64.0) | 5527 (65.7) | 0.011 |
| Heart failure | 1841 (34.4) | 881 (28.7) | 2722 (32.4) | <0.001 |
| CKD | 741 (13.9) | 275 (9.0) | 1016 (12.1) | <0.001 |
| Hyperlipidemia | 3619 (67.7) | 2121 (69.2) | 5740 (68.3) | 0.175 |
| STEMI<br>Anterior/Anterolateral | 1622 | 902 | 2524 |  |
| STEMI<br>Inferior/Inferolateral<br>Inferoposterior | 1371 | 967 | 2338 |  |
| STEMI lateral | 114 (2.1%) | 93 (3.0%) | 207 (2.5%) | 0.0130 |
| STEMI posterior | 52 (1.0%) | 24 (0.8%) | 76 (0.9%) | 0.4430 |
| STEMI<br>unspecified/multiple | 2185 | 1080 | 3265 |  |

CKD – chronic kidney disease; STEMI –ST elevation myocardial infarction; Trop-Troponin

### Mortality

At 1 year, the survival rate was 84% in the High trop group and 92.9% in the Low trop group (p<0.001). By 5 years, survival decreased to 75% in the High trop group and 85.7% in the Low trop group (p<0.001), demonstrating a substantial and persistent difference in long-term outcomes between the groups, as shown in Figure 1 and Table 2.

**Table 2.** Survival rate.

CKD – chronic kidney disease; STEMI –ST elevation myocardial infarction; Trop-Troponin
| Table 2 – Survival rate |  |  |  |
| --- | --- | --- | --- |
| Time after hospitalization | Trop > Rule In, No. (%)<br>n=5330 | Trop < Rule In, No. (%)<br>n=3064 | p Value |
| 1 year | 4477 (84%) | 2845 (92.9%) | <0.001 |
| 5 years | 3999 (75%) | 2626 (85.7%) | <0.001 |

**Figure 1.**
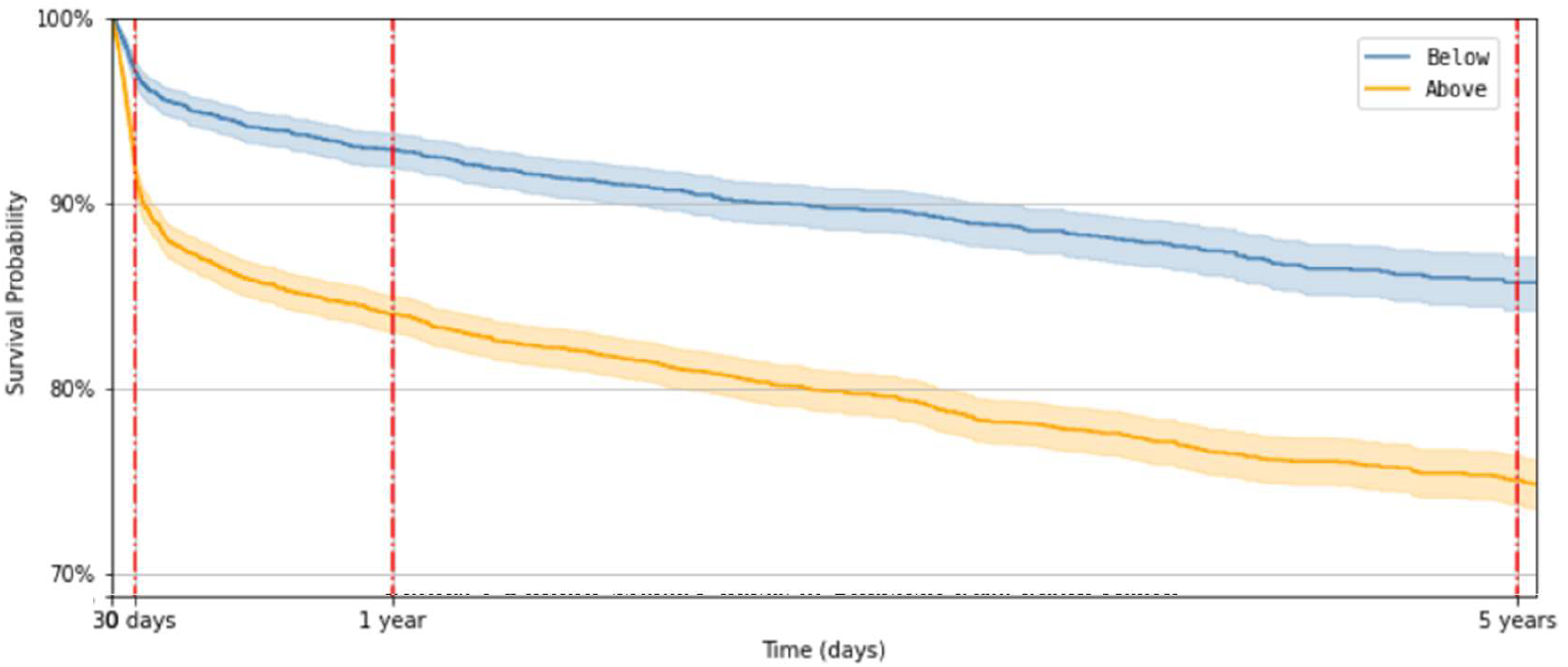
Kaplan-Meier survival curves of STEMI patients with Low troponin on admission (blue lines) and elevated troponin (orange line), including 95% CIs. The graph demonstrates a significantly better, early, and long-term survival of patients with Low troponin on admission. The 1-year (92.9%) and 5-year (85.7%; n=3064) survival rates were significantly higher in the low range troponin group compared with the elevated troponin group (84% and 75%, respectively; n=5330; P<0.001)

### Univariate and Multivariate Analysis of 5-Year Mortality

A univariate and a multivariate Cox regression model were applied to identify independent predictors of 5-year mortality following STEMI. As shown in Table 3 and in Figure 2, after adjustment for clinically relevant covariates, the initial hs-cTn level was strongly associated with increased risk of death (HR 1.15, 95% CI 1.14-1.16, p<0.001). Additional predictors included CKD (HR 1.37, 95% CI 1.35-1.39, p<0.001), age above 75 (HR 1.25, 95% CI 1.24-1.26, p<0.001), and female sex (HR 1.25, 95% CI 1.23-1.26, p<0.001). Diabetes mellitus (HR 1.03, 95% CI 1.02-1.04, p<0.001) and hypertension (HR 1.03, 95% CI 1.02-1.04, p<0.001) were also independently associated with increased mortality, but with a smaller effect size. A history of heart failure was not a significant predictor in the adjusted model. Overall, several variables showed attenuation of effect sizes relative to univariate analyses, while the initial hs-cTn level remained a robust independent predictor.

**Table 3.** Uni- vs Multi-Variate – Cox regression.

| Table 3– Uni- vs Multi- Variate – Cox regression |  |  |  |  |
| --- | --- | --- | --- | --- |
|  | Univariate (HR; 95% CI) |  | Multivariate (HR; 95% CI) |  |
|  | Predictor | P value | Predictor | P value |
| CKD | 1.56 (1.54-1.58) | <0.001 | 1.37 (1.35-.1.39) | <0.001 |
| Age above 75 | 1.43 (1.41-1.44) | <0.001 | 1.25 (1.24-1.26) | <0.001 |
| Gender (female) | 1.39 (1.38-1.40) | <0.001 | 1.25 (1.23-1.26) | <0.001 |
| Trop Rule-In | 1.19 (1.18-1.20) | <0.001 | 1.15 (1.14-1.16) | <0.001 |
| Diabetes | 1.16 (1.15-1.17) | <0.001 | 1.03 (1.02-1.04) | <0.001 |
| Hypertension | 1.19 (1.18-1.21) | <0.001 | 1.03 (1.02-1.04) | <0.001 |
| Heart failure | 1.17 (1.15-1.18) | <0.001 | 1.01 (1-1.2) | 0.19 |

CKD – chronic kidney disease

**Figure 2.**
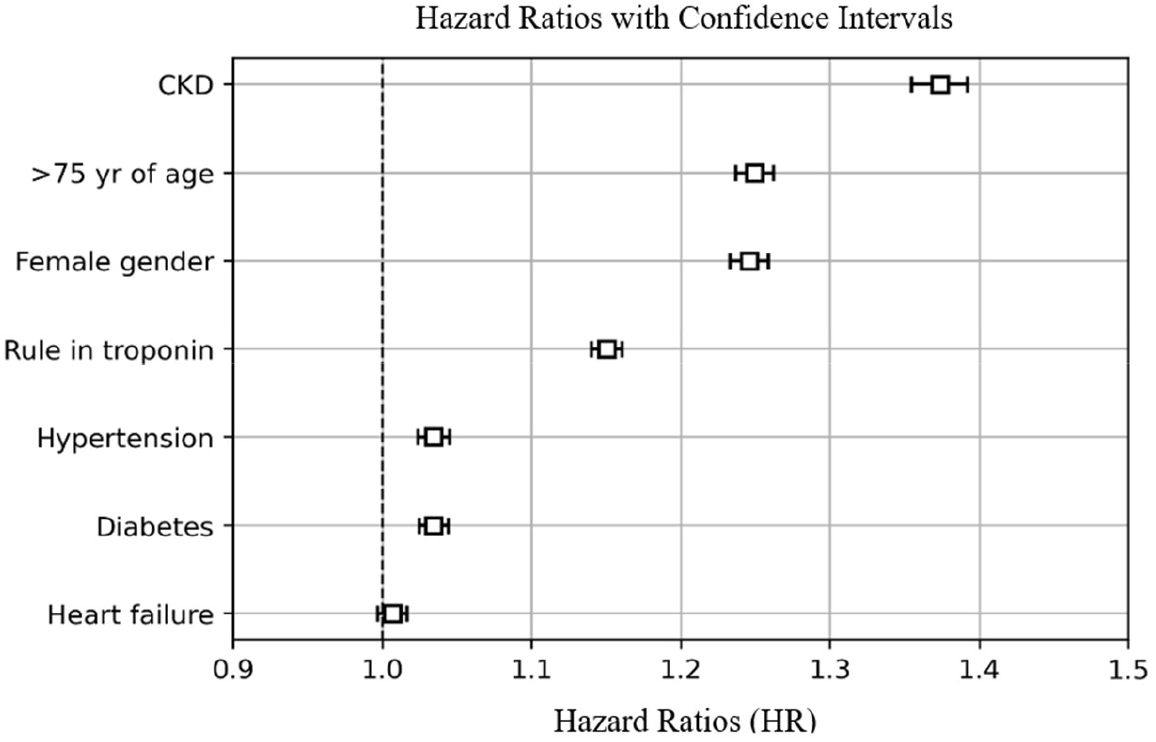
Cox regression model of possible confounders in STEMI patients. The model predicts significantly poorer 5-year survival in patients with elevated troponin on admission, with an adjusted hazard ratio of 1.15 (1.14-1.16; P<0.01). As expected, other variables such as CKD [adj HR 1.37 (1.35-1.39); P<0.01], female gender [adj HR 1.25 (1.23-1.26); P<0.01], and age above 75 [adj HR 1.25 (1.24-1.26); P<0.01], had a significantly higher long-term 5-year mortality.

## Discussion

This study is one of the largest-scale studies to evaluate the prognostic value of initial troponin on the long-term prognosis of STEMI patients.

Our initial hypothesis was that STEMI patients with normal troponin levels on arrival to the hospital have unique characteristics with favorable outcomes and may be regarded as a separate entity. Our main findings are: 1) STEMI patients treated with primary PCI who had an initial troponin level below the ESC rule-in threshold had an improved short- and long-term survival compared to patients presenting with elevated troponin. 2) After comprehensive multivariable adjustment, admission troponin remained a robust independent predictor of 5-year mortality, alongside established risk factors including age >75 years, female sex, and CKD.

Our results align with previous smaller studies showing the effect of initial troponin on the prognosis of STEMI patients^6,7^. This association persists even after adjustment for age and major comorbidities, supporting the independent prognostic significance of initial troponin. Unlike earlier studies that focused mainly on short-term outcomes^6^, or a smaller sample size^7,8^, our study shows a lasting impact of initial troponin on long-term mortality. We achieved this by analyzing a substantially larger cohort and using a comprehensive multivariable model that includes detailed comorbidity data from a database with up to 8 years of follow-up.

### Prognostic Significance of Admission Troponin in STEMI

More than one-third of our cohort presented with low troponin levels on arrival. These patients had significantly improved survival at 5 years compared with patients with elevated troponin on admission. Our findings align with and extend previous observations regarding the prognostic value of admission troponin in STEMI patients undergoing primary PCI.

Wanamaker et al. reported that 47.2% of STEMI patients had undetectable or normal troponin on presentation, with significantly lower in-hospital mortality compared to those with elevated levels (1.8% vs. 5.1%)^6^. Similarly, Coelho-Lima et al. demonstrated that high admission hs-cTnt levels of >515 ng/L independently predicted long-term mortality (HR 1.27)^8^. Our study extends these observations to a larger cohort with longer follow-up, demonstrating that the prognostic impact of admission troponin persists for at least 5 years post-event, even with a modest hs-cTnt at admission. In our study, elevated admission troponin was independently associated with mortality (HR 1.15, 95% CI 1.14-1.16). Unlike previous studies that used 5-10 times above the rule-in initial troponin levels as the cut-off for high mortality^6,8^, our study suggests that even a modest increase in admission hs-cTn levels affects short- and long-term mortality after STEMI. Even after adjusting for multiple covariates, admission troponin retained its predictive value, suggesting that it captures information not fully explained by traditional risk factors.

### Mechanisms Underlying Normal Troponin at STEMI Presentation

The substantial proportion of STEMI patients presenting with troponin below the rule-in threshold likely represents three distinct clinical phenotypes. First, patients arriving within the earliest phase of symptom onset may present before sufficient troponin release has occurred. Troponin elevation typically begins within 1-4 hours of symptom onset^5^, with a significant portion of patients with troponin below the rule-in threshold within the first 2 hours of symptom onset^9^. Second, this cohort may include patients with incomplete coronary occlusion or those who achieved spontaneous or pharmacological reperfusion prior to hospital arrival, resulting in smaller territories of myocardial necrosis despite characteristic ECG changes^10^. The third mechanism may involve patients with ischemic preconditioning and a well-established collateral coronary system, which may also result in a smaller territory of myocardial injury or necrosis.

### Baseline Characteristics and Comorbidities

The demographic and clinical differences between high and low troponin groups provide an important context for interpreting the mortality differences. The high troponin group was older and had a higher prevalence of multiple comorbidities known to independently predict mortality after STEMI^11^. The higher comorbidity burden in the high troponin group may reflect both increased vulnerability to larger infarcts and reduced physiologic reserve to tolerate myocardial injury or other major illnesses. Our results demonstrate several other risk factors to be independent predictors of mortality, the prominent ones are CKD, age above 75, and female gender. Interestingly, chronic kidney disease remained a powerful independent predictor with an HR of 1.37, consistent with extensive literature demonstrating that renal dysfunction is among the strongest predictors of mortality after STEMI^12^.

Many studies have found gender disparities in STEMI outcomes, with significantly higher mortality in women^13,14^. Some studies suggest this gap can be attributed to the increase in comorbidities in female STEMI patients, such as older age, and delay in treatment^13^, while others found a significant gap even after adjustment for co-morbidities^14,15^. In our study, even after adjustment, female gender remained an independent risk factor for mortality with an HR of 1.25.

### Implications for Risk Stratification

Our findings support the incorporation of admission troponin into risk stratification algorithms for STEMI patients undergoing primary PCI. While current risk scores, such as the GRACE and TIMI risk scores, incorporate various clinical and laboratory parameters, admission troponin is not universally included. Our data and others suggest that even in the presence of ST-elevation, the magnitude of initial troponin elevation provides independent prognostic information that could enhance risk prediction models. In the future, initial troponin and other risk factors may be classified as a new entity, “low risk STEMI” with different evaluation, personalized prognosis, and management, including shorter length of hospitalization, intensity of follow-up, and secondary prevention strategies.

### Limitations

Several limitations warrant consideration. First, the retrospective design limits our ability to establish causality and may introduce selection bias. Second, unfortunately, we lacked data on symptom-to-door time for all patients, which would provide important context for interpreting admission troponin levels. Patients presenting very early may have low troponin levels simply due to insufficient time for biomarker release, rather than smaller infarcts. Third, we used different hs-cTn assays across centers (Roche hs-cTnt and Abbott hs-cTni), though we applied assay-specific ESC rule-in thresholds to maintain consistency. Fourth, we lacked imaging data, such as cardiac MRI or echocardiographic parameters of left ventricular function, for the majority of patients, which would have allowed direct correlation between admission troponin and infarct size. Finally, cause-specific mortality data were not available, limiting our ability to determine whether the observed mortality differences were driven by cardiac versus noncardiac deaths. However, given the nature of our database, it likely presents the “real world”.

## Conclusions

In this large multicenter cohort of STEMI patients, more than one-third presented with troponin levels below the ESC rule-in threshold and demonstrated significantly better short- and long-term survival compared to those with elevated admission troponin. After comprehensive adjustment for demographic and clinical variables, admission troponin remained a powerful independent predictor of 5-year mortality. These findings suggest that initial troponin may reflect critical information about the extent of myocardial injury and ischemic burden that translates into sustained prognostic implications. Admission troponin should be considered an important component of risk stratification in STEMI patients, with potential implications for tailoring post-discharge management and follow-up intensity.

## Data Availability

The data that support the findings of this study are available upon request from the corresponding author. The data are not publicly available due to ethical restrictions and participant privacy regulations imposed by the Helsinki Committee

## Notes

### Competing Interest Statement

The authors have declared no competing interest.

### Clinical Trial

This is a retrospective study and is not required to an international trial registry

### Author Declarations

Tzafon Medical Center Helsinki Committee

